# Central inhibition of pain is augmented in women with self-injurious behavior

**DOI:** 10.1101/2021.05.12.21257091

**Authors:** Maria Lalouni, Jens Fust, Johan Bjureberg, Granit Kastrati, Robin Fondberg, Peter Fransson, Nitya Jayaram-Lindström, Eva Kosek, Clara Hellner, Karin B. Jensen

**Affiliations:** Department of Clinical Neuroscience, Karolinska Institutet, Stockholm, Sweden; Centre for Psychiatry Research, Karolinska Institutet & Stockholm Health Care Services, Stockholm County Council, Stockholm, Sweden; Stanford University, Department of Psychology, Stanford, California, USA; Uppsala University, Department of Surgical Sciences, Uppsala, Sweden

**Author notes:** Corresponding author: Jens Fust. Shared first authorship (Maria Lalouni, Jens Fust).

## Abstract

Individuals who engage in nonsuicidal self-injury (NSSI) have demonstrated higher pain thresholds and tolerance compared with individuals without NSSI. The objective of the study was to assess which aspects of the pain regulatory system that account for this augmented pain perception. In a cross-sectional design, 81 women, aged 18-35 (mean [SD] age, 23.4 [3.9]), were included (41 with NSSI and 40 healthy controls). A quantitative sensory testing protocol, including heat pain thresholds, heat pain tolerance, pressure pain thresholds, conditioned pain modulation (assessing central down-regulation of pain), and temporal summation (assessing facilitation of pain signals) was used. Thermal pain stimuli were assessed during fMRI scanning and NSSI behaviors and clinical symptoms were self-assessed. NSSI participants demonstrated higher pain thresholds during heat and pressure pain compared to controls. During conditioned pain modulation, NSSI participants showed a more effective central down-regulation of pain for NSSI participants. Temporal summation did not differ between the groups. There were no correlations between pain outcomes and NSSI behaviors or clinical characteristics. The fMRI analyses revealed increased activity in the primary and secondary somatosensory cortex in NSSI participants, compared to healthy controls, which are brain regions implicated in sensory aspects of pain processing. The findings suggest segregated inhibitory mechanisms for pain and emotion in NSSI, as pain insensitivity was linked to enhanced inhibitory control of pain in spite of significant impairments in emotion regulation. This may represent an endophenotype associated with a greater risk for developing self-injurious behavior.

---

The sensation of pain is inherently aversive and shapes us to avoid situations that cause harm to our bodies. Yet, there are individuals who deliberately induce pain by self-injury. Nonsuicidal self-injury (NSSI) is defined as intentional self-inflicted tissue damage, performed without suicidal intent, and for purposes that are not socially sanctioned.^1^ NSSI is highly prevalent in adolescents and young adults^2^ and is associated with high psychiatric comorbidity^3,4^ and an increased risk for suicide.^5,6^

Even though NSSI results in tissue damage, the majority of individuals who self-injure report that they feel little or no pain while they hurt themselves.^7-9^ Insensitivity to pain may be a risk factor for developing NSSI and individuals who do not experience physical pain during NSSI have shown to have a doubled incidence of suicide attempts, compared with individuals who experience pain during NSSI.^10^ The altered sensitivity to pain found in individuals with NSSI is not limited to self-injurious behaviors. Laboratory studies demonstrate that participants with NSSI display heightened pain thresholds and tolerance.^11,12^ Borderline symptomatology,^11,13^ self-critical cognitive style,^14,15^ and dysregulation of the opioid system^16^ have been proposed as underlying causes to the pain insensitivity, but results are inconclusive. One strategy to shed light on why individuals who engage in NSSI feel less pain is to investigate their pain system.

An individual’s pain modulation profile can be assessed by combining two psychophysical pain testing protocols: conditioned pain modulation and temporal summation,^17^ assessing central down regulation of pain and facilitating of pain signals, respectively. Both conditioned pain modulation and temporal summation have been suggested as biomarkers for chronic pain.^18,19^ Pain regulation can also be assessed with functional magnetic resonance imaging (fMRI), which has the potential to elucidate the brain regions involved in pain regulation.

The aim of the present study was to characterize the pain regulation in young women with NSSI and to contrast this with a matched sample of healthy women (HC). A further aim was to assess whether specific aspects of pain regulation would correlate with the participants’ self-reported NSSI behaviors (eg, frequency or duration of self-injurious behavior) and clinical characteristics (eg, borderline symptoms or self-criticism). Also, we wanted to determine the neural correlates of experimental pain in participants with NSSI compared with HC.

## Methods

This cross-sectional study was approved by the Regional Ethical Review Board in Stockholm (2018/1367-31/1) and pre-registered on the Open Science Framework May 27^th^ 2019 (osf.io/gujwt). The fMRI analysis was explorative and therefore not preregistered.

### Setting

All study visits took place at the MR Center, Karolinska University Hospital in Stockholm, Sweden, between May 2019 and August 2020, with a brief disruption during the first wave of the Covid-19 outbreak from mid-March to the end of May 2020.

### Participants

Based on a recent meta-analysis, we estimated that we needed 29 participants in each group to achieve 80% power to detect difference in pain threshold between NSSI participants and HC. We decided to recruit 40 participants to be able to detect the potentially smaller differences between groups in the other pain tests. Participants with NSSI (n=41) and HC (n=40) were recruited between April 2019 and June 2020, through flyers in waiting rooms at outpatient psychiatric clinics (NSSI) and advertisements in social media (NSSI and HC). The advertisement for HC was adjusted during the inclusion procedure so that age and educational level would match those of the NSSI participant. General inclusion criteria were: a) woman, b) 18-35 years, and c) right-handed. General exclusion criteria were: d) chronic inflammatory, auto-immune, or other somatic disorder requiring treatment, e) pain condition, f) contraindication for fMRI (eg, metal implant, pregnancy, claustrophobia), g) suicide attempts during the last year, h) suicidal plans or acute risk for suicide. Specific inclusion criteria for participants with NSSI: i) self-injury ≥ 5 days during the last year. Specific exclusion criteria for HC: j) treatment for depression or anxiety. Participants with NSSI received a remuneration of $171 (1500 SEK) and HC received $137 (1200 SEK).

### Measures and equipment

#### The pain testing included

1. *Heat pain thresholds and tolerance* were assessed with a thermal stimulator (MSA Thermal Stimulator Somedic, Hörby, Sweden) using a (30×30 mm) thermode.
2. *Pressure pain thresholds* were assessed with a handheld algometer (Somedic Algometer version II, Hörby, Sweden) with a 1 cm^2^ round rubber tip. The algometer records pressure in kilopascals (kPa) and was set to indicate when a speed of 50 kPa/s was used. An automatic pressure apparatus (APA Somedic, Hörby, Sweden), with identical characteristics as the algometer, was used for 5 persons in the NSSI group. This apparatus was later replaced with the algometer, due to technical difficulties.
3. *Conditioned pain modulation*. Pressure pain thresholds (kPa) applied with an algometer on the left calf was used as test stimulus and ischemic pain to the lower right arm was used as conditioning stimulus. The ischemic pain was induced with a blood pressure cuff connected to a cuff inflation system (Hokanson Inc, Bellevue, WA, USA) in combination with a handheld dumbbell (1 kg) that was flexed up and down (1 movement per second).
4. *Temporal summation* was assessed with PinPrick Stimulators (MRC Systems, Heidelberg, Germany). The PinPrick kit consists of seven stimulators, each with a flat contact area of 0.25 mm in diameter and weights between 8 mN and 512 mN. A stimulator that induced low pain, between 1/10 and 3/10 on a numerical rating scale (NRS), was individually tried out and subsequently used.

### fMRI

Magnetic resonance images were acquired with a 3T General Electric 750 MR scanner. Functional scans using T2*-weighted single-shot gradient echo planar imaging were collected, using the following parameters: repetition time/ echo time = 2000/30 ms, flip angle = 70°, field of view = 220 × 220 mm, matrix size = 72 × 72, 42 slices, slice thickness = 3 mm with a 0.5-mm gap, interleaved slice acquisition. Anatomical images were acquired with a high-resolution BRAVO 3D T1-weighted image sequence (1 × 1 × 1-mm voxel size, 176 slices).

### Questionnaires

Frequency and function of NSSI were assessed with the Functional Assessment of Self-Mutilation (FASM).^8,9^ Borderline symptoms were assessed with the short version of the Borderline Symptom List (BSL-23).^20^ The Self-Rating Scale (SRS) was used to assess self-criticism.^21^ Emotion regulation was assessed with the brief 16 item version of the Difficulties with Emotion Regulation Scale (DERS-16).^22^ Suicidal behaviors were self-assessed online with the Self-Injurious Thoughts and Behaviors Interview-Short Form-Self Report (SITBI-SF-SR).^9,23^ NSSI frequency was measured by the question in FASM: “How many days have you engaged in the type of self-harm mentioned in the previous questions?”. The participants were asked to check one of four boxes: 1-4, 5-15, 15-50, 50-. Severe NSSI methods score was calculated by summing up the prevalence of self-reported use of severe NSSI methods in FASM (cutting/carving, burning, scraping, and erasing the skin), as defined by Lloyd-Richardson et al.^8^ NSSI recency was measured on visit 1 by the question: “How many days have passed since you last engaged in self-harm behavior?” NSSI duration was measured by a question from FASM: “How old were you when you first harmed yourself in this way [as described by the previous questions in FASM]?”

### Study procedure

Participants gave informed consent to participate and completed all self-report questionnaires through a secure online data collection platform (BASS, eHealth Core Facility, Karolinska Institutet, Sweden). NSSI participants met a licensed psychologist once who assessed their potential risk of participating in the study and asked about their current psychiatric comorbidity. Pain tests were then assessed at two consecutive visits for both NSSI participants and HC. Participants were asked to withhold from as needed medication or drugs that could affect their pain regulation (eg, tranquilizers, NSAIDs, paracetamol) 24 hours before the pain tests. However, if they urgently needed to take such medication, they could do so.

#### Visit 1 - Behavioral pain tests

Heat pain threshold and tolerance were assessed with a thermode on the participants left calf using repeated heat stimuli (5 seconds long with an interstimulus interval of 30 seconds). Pain was rated on a continuous 0-10 scale (NRS). The participants were exposed to between 13-24 heat stimulations (depending on the pain sensitivity of the participant). The temperature never exceeded 50°C, to avoid the risk of tissue damage (for a detailed description of the heat pain calibration procedure, see Supplementary A). Pressure pain thresholds were assessed with an algometer on the participants’ anterior, distal part of the left thigh (quadriceps femoris). Participants indicated when the pain threshold was reached by pressing a button. A total of 3 pressure stimulations were administered. During conditioned pain modulation pressure pain thresholds were assessed on the participants’ left calf twice before the conditioning stimulus was induced, twice during the conditioning stimulus, and twice after the conditioning stimulus was removed (Figure 1A). The last two pressure pain thresholds were assessed to ensure that the participants’ pain sensitivity returned to baseline.

**Figure 1.**
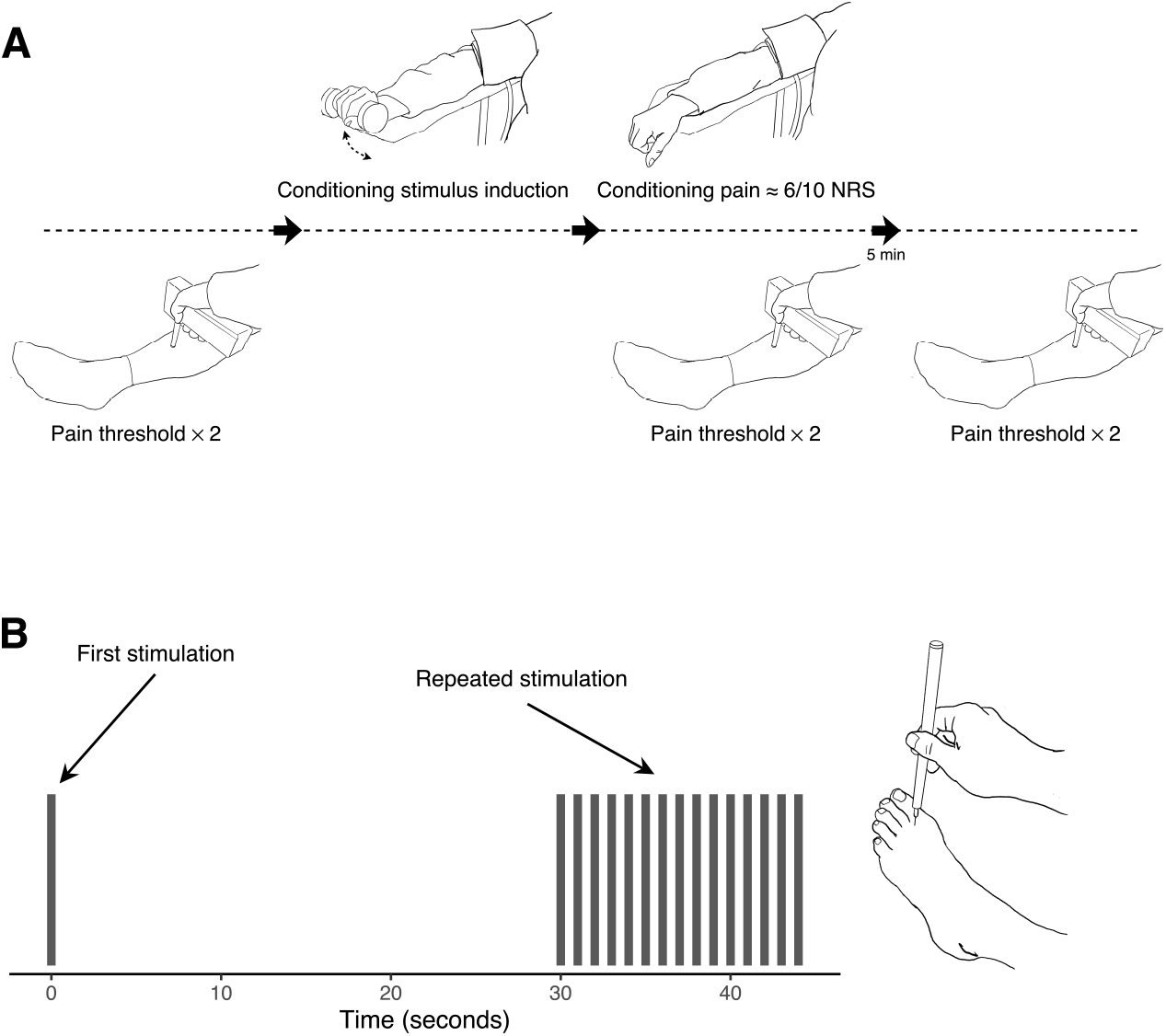
Testing procedure for conditioned pain modulation (A) and temporal summation (B). Abbreviation: NRS, numerical rating scale.

#### Visit 2 - Pain tests in the MR-scanner

During temporal summation, pain was induced with a PinPrick stimulator on the participants’ left foot, between the big toe and second toe (Figure 1B). Pain was induced a) once, and b) 15 times (once per second), four times each with a break of 30 seconds between the four trials. Imaging data for temporal summation will be presented elsewhere. Individually calibrated heat pain (approximately NRS 5/10) was used to provide 30 second blocks of “pain on” (three blocks) and “pain off” (three blocks). The heat pain was induced on the participants’ left calf with a thermode. Each block was preceded by a visual cue on a screen inside the MR-scanner (10 s duration) indicating if the block would include pain or not. After each stimulation the pain was rated on a 0-10 NRS.

### Statistical Analysis

#### Behavioral data

Baseline variables for the HC and NSSI groups were compared using χ^2^ for categorical variables and 2-tailed *t* tests for continuous variables. Pressure pain thresholds were defined as the mean kPa of three trials. Heat pain threshold and heat pain tolerance were calculated by fitting a linear regression to each participant’s pain ratings, with stimulus temperature as a predictor. Heat pain threshold was defined as the participant’s predicted NRS 1/10 and heat pain tolerance was defined as the participants predicted NRS 6/10. Comparisons between the two groups’ pain threshold and tolerance were made using Wilcoxon rank sum test. The conditioned pain modulation effect was defined as the difference between pressure pain thresholds, before and during the conditioning stimulus. A mixed effect model was fitted with conditioning stimulus (on/off), group (NSSI/HC) and the interaction between conditioning stimulus and group specified as fixed effects. As random effects, we used by-subject intercepts and by-subject slopes for conditioned stimulus. Temporal summation was assessed by the wind-up effect, defined as the difference between the first pain rating and the maximum pain rating during the repeated stimulation. A mixed effects model, in which stimulus (first/max), group (NSSI/HC), and the interaction between stimulus and group was specified as fixed effects, and by-subject intercepts and by-subject slopes for stimulus was specified as random effects. Lastly, pain outcomes were correlated with self-report data of NSSI behavior and clinical characteristics using Kendall rank correlation coefficient. Because of the exploratory nature of these correlations, we did not correct for multiple comparisons. The statistical analyses were conducted in R^24^ and Stata.^25^ Mixed effect model was fitted R packages lme4^26^ and lmerTest.^27^

#### Neuroimaging data

Data analyses were performed using Statistical Parametric Mapping 12 (SPM12)^28^, and Matlab2014 (The MathWorks, Inc, Natick, MA). For preprocessing parameters, see Supplement A. A first-level general linear model (GLM) was built for each individual and included the following regressors of interest: pain cue, pain-on, pain rating, no-pain cue, no-pain. Regressors of interest were convolved with the canonical hemodynamic response function. Six motion parameters were added as regressors of no interest. On the group level, one-sample t-tests were performed to determine brain activation patterns across all participants. Two-sample t-tests were used to determine differences in brain activation between groups. Statistical significance was considered at an initial statistical threshold of P<.05 FWE corrected over the entire brain, and then cluster-level P<.05 FEW corrected.

## Results

A total of 81 women, mean [SD] age 23.4 [3.9], were included in the study (Table 1). Because of technical difficulties and problems with removal of metal objects (eg, piercings) only 31 of 41 NSSI participants were scanned and included in the fMRI analysis. For participants with NSSI, the age of self-injury onset was mean [SD] 13.2 [3.1] years. Of the 41 participants with NSSI, 28 regularly used medication, compared with 4 in HC. Most commonly used medication reported by participants with NSSI was selective serotonin reuptake inhibitors (SSRIs) used by 19 of the 41 participants. During the clinical interview 32 out of 39 replied that they had a psychiatric diagnosis. For two participants the response is missing. The most frequently reported diagnosis was depression (n=21), followed by anxiety disorder (n=12), and borderline personality disorder (n=11). Most common types of self-harm reported for the last year were hitting oneself on purpose (n=32) and cutting or carving on the skin (n=31). Mean number of methods used were [SD] 5.6 [2.2], range 2-12. The most commonly reported reasons for self-harm were “to punish yourself” and “to stop bad feelings”.

**Table 1.** Demographic and clinical characteristics

| Characteristic | Mean (SD) |  |  | Statistic | P value |
| --- | --- | --- | --- | --- | --- |
|  | NSSI | HC | Total |  |  |
| Number of participants | 41 | 40 | 81 |  |  |
| Demographic characteristics |  |  |  |  |  |
| Age, mean (SD), years | 22.7 (3.6) | 24.0 (4.1) | 23.4 (3.9) | $t = -1.54$ | .128 |
| Educational level, n (%) |  |  |  |  |  |
| Elementary school | 7 (17.1) | 7 (17.5) | 14 (17.3) | $\chi^2 = 1.19$ | .755 |
| High school | 27 (65.9) | 24 (60.0) | 51 (63.0) |  |  |
| College degree | 5 (12.2) | 8 (20.0) | 13 (16.1) |  |  |
| Other | 2 (4.9) | 1 (2.5) | 3 (3.7) |  |  |
| Clinical characteristics |  |  |  |  |  |
| BSL-23, mean (SD) | 2.3 (0.9) | 0.4 (0.3) | 1.3 (1.2) | $t = -13.0$ | <.001 |
| SRS, mean (SD) | 33.7 (8.5) | 16.2 (7.9) | 25.0 (12.0) | $t = -9.7$ | <.001 |
| DERS-16, mean (SD) | 56.2 (13.3) | 27.1 (9.9) | 41.8 (18.8) | $t = -11.2$ | <.001 |

Abbreviations: SD, standard deviation; NSSI, nonsuicidal self-injury; HC, healthy control participants; BSL-23, Borderline Symptom List; SRS, Self-Rating Scale (self-criticism); DERS-16, 16 Item Version of the Difficulties in Emotion Regulation Scale.

### 1. Heat pain thresholds and tolerance

Model estimated heat pain threshold [SD] was 46.3 [2.9] °C for the NSSI group and 44.8 [2.1] °C for the HC group (Figure 2A). There was a significant difference between the groups, *P* = .031. Model estimated heat pain tolerance [SD] was 49.3 [2.4] °C for the NSSI group and 48.5 [1.0] °C for the HC group (Figure 2A). There was no significant difference between the groups, *P* = .192.

**Figure 2.**
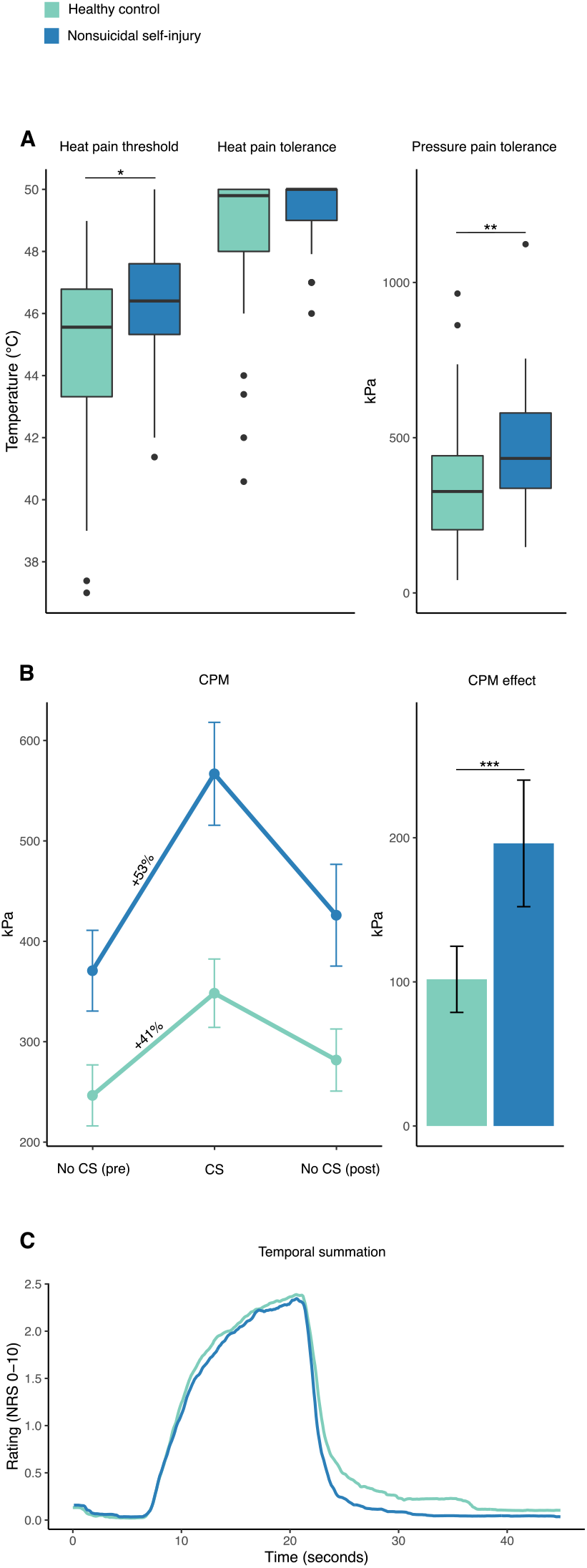
Results from pain testing. (A) Boxplots (including median, interquartile range, and upper and lower adjacent values) of heat pain threshold, heat pain tolerance, and pressure pain threshold. (B) Mean pressure pain threshold during CPM procedure and CPM effect (error bars represent 95% confidence interval). (C) Mean pain ratings during repeated stimulation in the temporal summation procedure. Abbreviations: CPM, conditioned pain modulation; CS, conditioning stimulus. * = *P* < .05, ** = *P* < .01, *** = *P* < .001.

**Figure 3.**
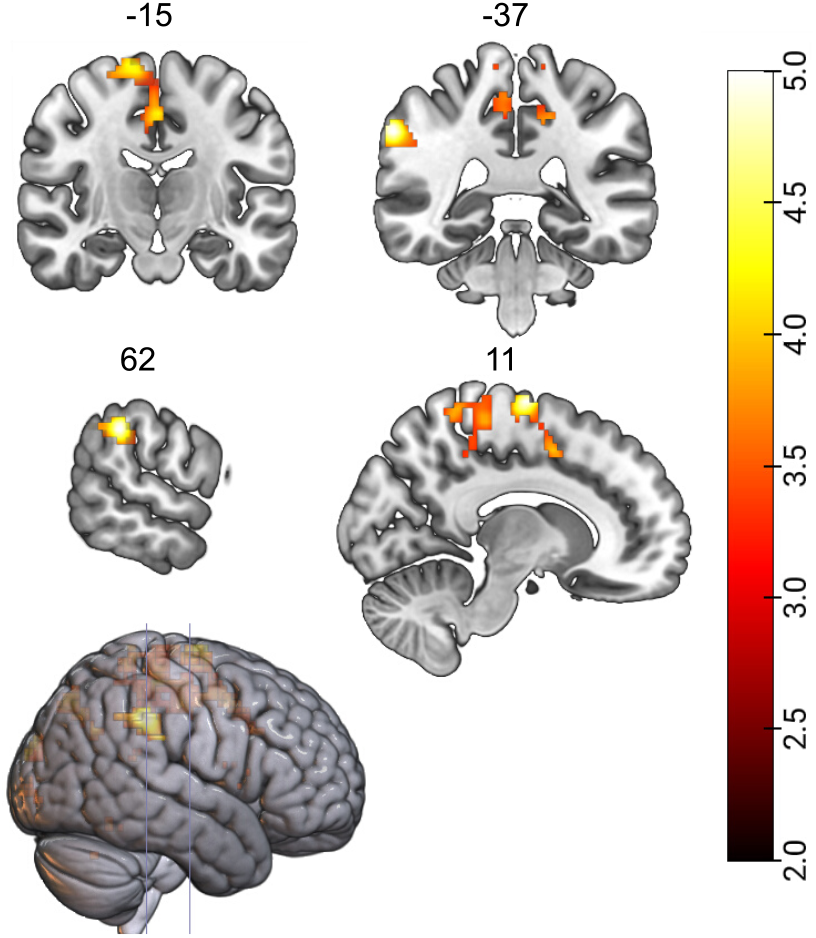
Increases in brain activation for non-suicidal self-injury participants (NSSI) patients compared to controls (HC) during tonic, subjectively calibrated heat pain. The primary and secondary somatosensory cortexes, contralateral to the leg stimulation, were more activated among NSSI individuals compared to HC. Anatomical locations of cluster-level peak activations (T-scores) are given in MNI-152 stereotactic atlas coordinates (top row: y-coordinates for coronal slices; bottom row: x-coordinates for sagittal slices) and labeled through the Automated Anatomical Labelling digital atlas in MRIcron.

### 2. Pressure pain

Mean [SD] pressure pain threshold was 463 [189] kPa for the NSSI group and 356 [202] kPa for the HC group (Figure 2B). There was a significant difference between the groups, *P* = .007.

### 3. Conditioned pain modulation (CPM)

Model estimated CPM response was 196 kPa for the NSSI group and 102 kPa for the HC group (Figure 2C-D). There was a significant difference between the groups, *P* < .001.

### 4. Temporal summation (TS)

Model estimated wind-up effect was NRS 1.9/10 for the NSSI group and NRS 1.8/10 for the HC group (Figure 2E). There was no significant difference between the groups, *P* = .723.

### 5. Pain-related brain activations

Mean [SD] pain rating was NRS 4.5/10 [1.8] for the NSSI group and NRS 4.6/10 [2.0] for the HC group. Mean temperature [SD] during heat pain stimulation was 47.2 [0.8] °C for the NSSI group and 46.3 [1.7] °C for the HC group. The contrast between NSSI and HC revealed increased neural activations in the leg area of the primary somatosensory cortex (S1) (MNI coordinates: x = 11, y = -10, z = 72; t = 5.05, *P* = .022) and secondary somatosensory cortex (S2) (MNI coordinates: x = 62, y = -37, z = 37; t = 5.11; *P* = .013), contralateral to the stimulation site, corrected for multiple comparisons. There were no regions where HC had greater neural activations than NSSI.

### 6. Correlational analyses between pain outcomes and self-report data

We found no correlations between pain outcomes and self-report data of NSSI behavior or clinical characteristics (Table 2).

**Table 2.** Correlations between pain measures and self-report data

|  | 1 | 2 | 3 | 4 | 5 | 6 | 7 | 8 | 9 | 10 |
| --- | --- | --- | --- | --- | --- | --- | --- | --- | --- | --- |
| 1. Pressure threshold | 1 |  |  |  |  |  |  |  |  |  |
| 2. Heat threshold | 0.1 | 1 |  |  |  |  |  |  |  |  |
| 3. CPM effect | 0.13 | 0.22* | 1 |  |  |  |  |  |  |  |
| 4. NSSI frequency | -0.19 | -0.23 | -0.05 | 1 |  |  |  |  |  |  |
| 5. Severe NSSI methods | -0.19 | 0.08 | -0.15 | 0.13 | 1 |  |  |  |  |  |
| 6. NSSI recency | 0.07 | 0.09 | -0.04 | -0.21 | -0.09 | 1 |  |  |  |  |
| 7. NSSI duration | 0.06 | 0.01 | -0.04 | -0.14 | -0.16 | -0.16 | 1 |  |  |  |
| 8. BSL | 0.03 | -0.15 | -0.08 | 0.15 | 0.09 | -0.06 | -0.01 | 1 |  |  |
| 9. SRS | -0.06 | -0.11 | 0 | 0.28* | 0.09 | -0.22* | -0.12 | 0.35** | 1 |  |
| 10. DERS-16 | -0.02 | -0.17 | -0.06 | 0.28* | 0.1 | -0.06 | -0.16 | 0.5*** | 0.38*** | 1 |

Abbreviations: CPM, conditioned pain modulation; NSSI, nonsuicidal self-injury; BSL, Borderline Symptom List; SRS, Self-Rating Scale (self-critical cognitive style); DERS-16, 16 Item Version of the Difficulties in Emotion Regulation Scale.
\* = $P < .05$ ; \*\* = $P < .01$ ; \*\*\* = $P < .001$ .

## Discussion

Results from the present pain experiments demonstrate that participants with NSSI were less sensitive to pain compared with HC. Despite a marked inability to regulate negative emotions, pain regulation among individuals with NSSI was characterized by an enhanced top-down regulation of pain, as indicated by conditioned pain modulation. Participants with NSSI had higher pain thresholds for both pressure and heat pain, compared with HC. It has previously been shown that individuals with NSSI are less sensitive to pain stimuli,^11,12^ yet the underlying mechanisms have remained unclear. The present findings represent the first indication of an augmented pain-specific inhibitory mechanism in individuals with NSSI, compared with HC. The neuroimaging results revealed differences in cerebral pain processing between NSSI and HC, where NSSI participants displayed stronger activity in somatosensory brain regions during noxious heat. Since the brain imaging results are based on perception-matched heat pain (with higher temperatures given in NSSI), the increased involvement of somatosensory cortices (but not affective-motivational circuitry and saliency) may represent a neural pain signature with less salience signaling that enables more effective descending pain inhibition.

Both repeated exposure to pain and an antinociceptive endophenotype has been proposed as explanations to the pain insensitivity in NSSI.^29^ In a study comparing patients with borderline personality disorder with ongoing and previous NSSI, the group with ongoing NSSI experienced higher pain thresholds^30^ and Magerl et al^31^ found a correlation between recency of self-injury behavior and pressure pain threshold. However, the only longitudinal study assessing pain in NSSI by Koenig et al^32^ showed no change in pain sensitivity over time. In the current study, we observed no correlations between the pain thresholds or the conditioned pain modulation effect and the self-assessed NSSI characteristics (frequency, recency, duration, or use of severe methods). Thus, our results are in line with those of Koenig et al^32^ and also of Glenn et al^33^ who found no correlations between NSSI duration, or frequency (last month/year/lifetime) and pain sensitivity. Well-powered longitudinal studies are needed to thoroughly examine whether repeated exposure to pain, an inherited sensitivity to pain, or a combination of both, can explain the antinociceptive pain profile seen in individuals with NSSI.

It has been suggested that borderline symptomatology^11,13^ and a self-critical cognitive style,^14,15^ links NSSI and pain insensitivity. In our data, we found no correlations between the pain assessments and borderline symptoms or self-critical cognitive style. Emotion regulation is another type of central regulation in which individuals with NSSI typically deviate from HC. The NSSI participants in our study displayed augmented pain inhibition, while their ability to regulate emotions was significantly impaired, in agreement with previous studies suggesting that pain and emotion regulation are partly distinct and uncorrelated systems.^34^

Participants were exposed to tonic heat stimuli, which were individually calibrated to yield medium pain intensity, while measuring BOLD activity in the brain. During the painful blocks of heat, participants with NSSI had increased activations in somatosensory brain regions compared to HC, including one small cluster in the leg area of the contralateral primary somatosensory cortex (S1), and one small cluster in the secondary somatosensory cortex (S2). As the temperatures given to the leg were higher in the NSSI group, a putative interpretation is that the increase in activity in S1 and S2 are merely reflective of this fact.^35^ Yet, the higher temperature intensity among NSSI participants was associated with greater neural activations in sensory brain circuits during noxious input, as opposed to brain circuits related to salience and motivational value. As negative affect is likely to worsen pain, the increased sensory activation may prevent fear-related amplification of pain and/or facilitate inhibition. Previous data among healthy volunteers show increased activity in S1 during increased attention to a painful stimulus, and attenuation of S1 during self-administered pain,^36^ which could potentially reflect the differences between NSSI and HC seen here. Self-generated pain has been proven more predictable, and lead to decreased pain via self-attenuation.^37^ In contrast to their usual self-induced pain, it is possible that NSSI participants react with higher attention to the other-induced pain, as in this study procedure, and thereby display relatively greater S1 activity.

Knowledge of what aspect of the pain regulation is augmented in patients with NSSI can be used to inform medical and behavioral treatment targets.^38^ Also, pain profiling of psychiatric patients may help identify individuals at risk of developing NSSI and suicidal behaviors.

A limitation of the study is the cross-sectional study design, which precludes any causal conclusions regarding NSSI exposure and pain characteristics. Another limitation is the lack of control for psychiatric comorbidity. However, it is not likely that the psychiatric burden accounted for the pain insensitivity in the participants with NSSI. In fact, a large study comparing the pain sensitivity of 735 patients with depressive disorder (the most common psychiatric comorbidity of the participants with NSSI in our study) with 456 HC conclude that patients with depression were more sensitive to pain.^39^ Another limitation is the ceiling effect for heat tolerance, impeding the comparison between the groups. Strengths of the current study include the careful application of well-established pain tests and a rigorous inclusion procedure.

## Conclusions

We found higher pain thresholds and more effective central pain modulation in participants with NSSI compared with HC. The findings suggest segregated inhibitory mechanisms for pain and emotion in NSSI and the effective central down-regulation of pain may represent an endophenotype associated with a greater risk for developing NSSI and suicidal behaviors.

## Supporting information

Supplementary B: Results

## Data Availability

Unfortunately, we do not have ethical permission to share the data openly. Data will be made available upon request.

## Acknowledgements

We thank all the participants of the study for their participation. We also thank the psychiatric units that helped us inform about the study and the patient organization SHEDO for their support.

## References

1 Klonsky ED, Muehlenkamp JJ. Self-injury: a research review for the practitioner. J Clin Psychol 2007; 63: 1045–1056.

2 Cipriano A, Cella S, Cotrufo P. Nonsuicidal Self-injury: a systematic review. Front Psychol 2017; 8: 305–14.

3 Ghinea D, Edinger A, Parzer P, Koenig J, Resch F, Kaess M. Non-suicidal self-injury disorder as a stand-alone diagnosis in a consecutive help-seeking sample of adolescents. J Affect Disord 2020; 274: 1122–1125.

4 Nitkowski D, Petermann F. [Non-suicidal self-injury and comorbid mental disorders: a review]. Fortschr Neurol Psychiatr 2011; 79: 9–20.

5 Hamza CA, Stewart SL, Willoughby T. Examining the link between nonsuicidal self-injury and suicidal behavior: a review of the literature and an integrated model. Clin Psychol Rev 2012; 32: 482– 495.

6 Klonsky ED, May AM, Glenn CR. The relationship between nonsuicidal self-injury and attempted suicide: converging evidence from four samples. J Abnorm Psychol 2013; 122: 231–237.

7 Izadi-Mazidi M, Yaghubi H, Mohammadkhani P, Hassanabadi H. Assessing the functions of non-suicidal self-injury: factor analysis of functional assessment of self-mutilation among adolescents. Iran J Psychiatry 2019; 14: 184–191.

8 Lloyd-Richardson EE, Perrine N, Dierker L, Kelley ML. Characteristics and functions of non-suicidal self-injury in a community sample of adolescents. Psychol Med 2007; 37: 1183–1192.

9 Zetterqvist M, Lundh L-G, Dahlstrom O, Svedin CG. Prevalence and function of non-suicidal self-injury (NSSI) in a community sample of adolescents, using suggested DSM-5 criteria for a potential NSSI disorder. J Abnorm Child Psychol 2013; 41: 759–773.

10 Nock MK, Joiner TEJ, Gordon KH, Lloyd-Richardson E, Prinstein MJ. Non-suicidal self-injury among adolescents: diagnostic correlates and relation to suicide attempts. Psychiatry Res 2006; 144: 65–72.

11 Koenig J, Thayer JF, Kaess M. A meta-analysis on pain sensitivity in self-injury. Psychol Med 2016; 46: 1597–1612.

12 Kirtley OJ, O’Carroll RE, O’Connor RC. Pain and self-harm: A systematic review. J Affect Disord 2016; 203: 347–363.

13 Bekrater-Bodmann R, Chung BY, Richter I, Wicking M, Foell J, Mancke F et al. Deficits in pain perception in borderline personality disorder: results from the thermal grill illusion. Pain 2015; 156: 2084–2092.

14 Hooley JM, St Germain SA. Nonsuicidal self-injury, pain, and self-criticism: does changing self-worth change pain endurance in people who engage in self-injury? Clin Psychol Sci 2014; 2: 297–305.

15 Fox KR, O’Sullivan IM, Wang SB, Hooley JM. Self-criticism impacts emotional responses to pain. Behav Ther 2019; 50: 410–420.

16 Stanley B, Sher L, Wilson S, Ekman R, Huang Y-Y, Mann JJ. Non-suicidal self-injurious behavior, endogenous opioids and monoamine neurotransmitters. J Affect Disord 2010; 124: 134–140.

17 Yarnitsky D, Granot M, Granovsky Y. Pain modulation profile and pain therapy: between pro- and antinociception. Pain 2014; 155: 663–665.

18 Kennedy DL, Kemp HI, Ridout D, Yarnitsky D, Rice ASC. Reliability of conditioned pain modulation: a systematic review. Pain 2016; 157: 2410–2419.

19 Gruener H, Zeilig G, Gaidukov E, Rachamim-Katz O, Ringler E, Blumen N et al. Biomarkers for predicting central neuropathic pain occurrence and severity after spinal cord injury: results of a long-term longitudinal study. Pain 2020; 161: 545–556.

20 Bohus M, Kleindienst N, Limberger MF, Stieglitz R-D, Domsalla M, Chapman AL et al. The short version of the Borderline Symptom List (BSL-23): development and initial data on psychometric properties. Psychopathology 2009; 42: 32–39.

21 Hooley JM, Ho DT, Slater J, Lockshin A. Pain perception and nonsuicidal self-injury: a laboratory investigation. Personal Disord 2010; 1: 170–179.

22 Bjureberg J, Ljótsson B, Tull MT, Hedman E, Sahlin H, Lundh L-G et al. Development and validation of a brief version of the difficulties in emotion regulation scale: the DERS-16. J Psychopathol Behav Assess 2016; 38: 284–296.

23 Nock MK, Holmberg EB, Photos VI, Michel BD. Self-injurious thoughts and behaviors interview: development, reliability, and validity in an adolescent sample. Psychol Assess 2007; 19: 309–317.

24 Team RC. A language and environment for statistical computing. Vienna, Austria: R Foundation for Statistical Computing; 2013. 2012.

25 Stata S. StataCorp. 2019. Stata Statistical Software: Release 16. College Station, TX: StataCorp LLC.

26 Bates D, Mächler M, Bolker BM, Walker SC. Fitting linear mixed-effects models using lme4. J Stat Softw 2015; 67. doi:10.18637/jss.v067.i01.

27 Kuznetsova A, Brockhoff PB, Christensen RH. lmerTest package: tests in linear mixed effects models. J Stat Softw 2017; 82: 1–26.

28 Friston KJ, Holmes AP, Worsley K, Poline J, Firth C, Frackowiak R. Statistical parametric maps in functional imaging: a general linear approach. Hum Brain Mapp 1995; 2: 189–210.

29 Nock MK. Self-injury. Annu Rev Clin Psychol 2010; 6: 339–363.

30 Ludäscher P, Greffrath W, Schmahl C, Kleindienst N, Kraus A, Baumgärtner U et al. A cross-sectional investigation of discontinuation of self-injury and normalizing pain perception in patients with borderline personality disorder. Acta Psychiatr Scand 2009; 120: 62–70.

31 Magerl W, Burkart D, Fernandez A, Schmidt LG, Treede R-D. Persistent antinociception through repeated self-injury in patients with borderline personality disorder. Pain 2012; 153: 575–584.

32 Koenig J, Rinnewitz L, Niederbäumer M, Strozyk T, Parzer P, Resch F et al. Longitudinal development of pain sensitivity in adolescent non-suicidal self-injury. J Psychiatr Res 2017; 89: 81– 84.

33 Glenn JJ, Michel BD, Franklin JC, Hooley JM, Nock MK. Pain analgesia among adolescent self-injurers. 2014; : 1–6.

34 Jensen KB, Petzke F, Carville S, Fransson P, Marcus H, Williams SCR et al. Anxiety and depressive symptoms in fibromyalgia are related to poor perception of health but not to pain sensitivity or cerebral processing of pain. Arthritis Rheum 2010; 62: 3488–3495.

35 Bingel U, Lorenz J, Glauche V, Knab R, Gläscher J, Weiller C et al. Somatotopic organization of human somatosensory cortices for pain: a single trial fMRI study. Neuroimage 2004; 23: 224–232.

36 Helmchen C, Mohr C, Erdmann C, Binkofski F, Büchel C. Neural activity related to self-versus externally generated painful stimuli reveals distinct differences in the lateral pain system in a parametric fMRI study. Hum Brain Mapp 2006; 27: 755–765.

37 Lalouni M, Fust J, Vadenmark Lundqvist V, Ehrsson HH, Kilteni K, Jensen KB. Predicting pain: differential pain thresholds during self-induced, externally induced, and imagined self-induced pressure pain. Pain 2021; 162: 1539–1544.

38 Yarnitsky D, Granot M, Nahman-Averbuch H, Khamaisi M, Granovsky Y. Conditioned pain modulation predicts duloxetine efficacy in painful diabetic neuropathy. Pain 2012; 153: 1193–1198.

39 Hermesdorf M, Berger K, Baune BT, Wellmann J, Ruscheweyh R, Wersching H. Pain sensitivity in patients with major depression: differential effect of pain sensitivity measures, somatic cofactors, and disease characteristics. J Pain 2016; 17: 606–616.

