## Supplementary A: Methods for "Central inhibition of pain is augmented in women with self-injurious behavior"

**Pain calibration**

During pain calibration were exposed to between 13-24 heat stimulations (depending on the pain sensitivity of the participant). Each heat stimulation lasted 5 seconds, with an interstimulus interval of 35 seconds. Participants used a trackball to rate their pain intensity on a 0-10 NRS that was displayed on a screen after each stimulation. The heat pain calibration included four blocks of stimulations. In the first block, the first stimulation was 38°C. The temperature was then raised 1°C for each subsequent stimulation, until the participants rated their pain above NRS 1/10. In the second block, participants were given four stimulations starting from -1°C of the last temperature given in first block. For example, if the first block ended with 45°C, the participants were given a stimulation sequence of 44°C, 45°C, 46°C, 47°C, in a randomized order. The third and the fourth block, the stimulation sequence in block were once again given in a randomized order, but depending on the participants’ rating in the previous block the overall temperature was modified. If the maximum pain rating during the previous block did not exceed NRS 3/10 the overall temperature of the next stimulation sequence was raised 3°C: [44°C, 45°C, 46°C, 47°C] ⇒ [47°C, 48°C, 49°C, 50°C]. If the maximum pain rating during the previous block exceeded NRS 6/10 the overall temperature of the next stimulation sequence was lowered 1°C: [44°C, 45°C, 46°C, 47°C] ⇒ [43°C, 44°C, 45°C, 46°C]. If the max maximum pain rating during previous block was between NRS 3/10 - NRS 6/10, the temperature was raised 1°C: [44°C, 45°C, 46°C, 47°C] ⇒ [45°C, 46°C, 47°C, 48°C]. The temperature never exceeded 50°C, to avoid the risk of tissue damage. Heat pain threshold and heat pain tolerance were calculated by fitting a linear regression to each participant’s pain ratings during the last three blocks of the pain calibration (see Statistical analysis: Behavioral data in main article).

**Neuroimaging data preprocessing**

Data quality was reviewed using MRIQC (https://mriqc.readthedocs.io/en/stable/ ). Frame-wise displacement was used to determine if there was excessive head motion from one volume to the next. No participants had to be excluded from the analyses, as none displayed a frame-wise displacement of >0.5 in >15% of the images. The fmriprep pipeline for data preprocessing was used (<https://fmriprep.org/>), including realignment, and normalization to Montreal Neurological Institute (MNI) space. Images were then spatially smoothed using 8-mm full-width half-maximum Gaussian kernel. Data analyses were performed using Statistical Parametric Mapping 12 (SPM12), and Matlab2014 (The MathWorks, Inc, Natick, MA).
