## Supplementary B: Results for "Central inhibition of pain is augmented in women with self-injurious behavior"

**Conditioned pain modulation**

In accordance with the preregistered mixed effects model, order was specified as a fixed effect and by-subject random effect (model1 in eTable 1). Because of the high correlation between the effect of order and condition, the model did not converge. For that reason, we decided to drop order as a fixed and random effect. The results of the second model (model2 in eTable 2), without order as fixed and random effect, was reported in the main article. Visual inspection of diagnostic plots (eFigure 1) of the second model suggested that the assumption of homoscedacity and normality had been violated, possibly due to the large variance in the CPM effect in the NSSI group, that on average displayed higher CPM effect. Therefore, we also compared the mean CPM effect (difference between pain threshold, before and during conditioned stimulus) of the two groups, using non-parametric Wilocoxon rank sum test. We found a significant difference between the groups, *P* < .001. The code that accompanies these results can be found in ‘cpm_results.R’ in the R-script folder on https://osf.io/gujwt/. We adapted eTable 2 and 3 from from Appendix 5 in Meteyard and Davies^1^.

eFigure 1. Conditioned pain modulation: diagnostic plots of mixed effects model (model2)
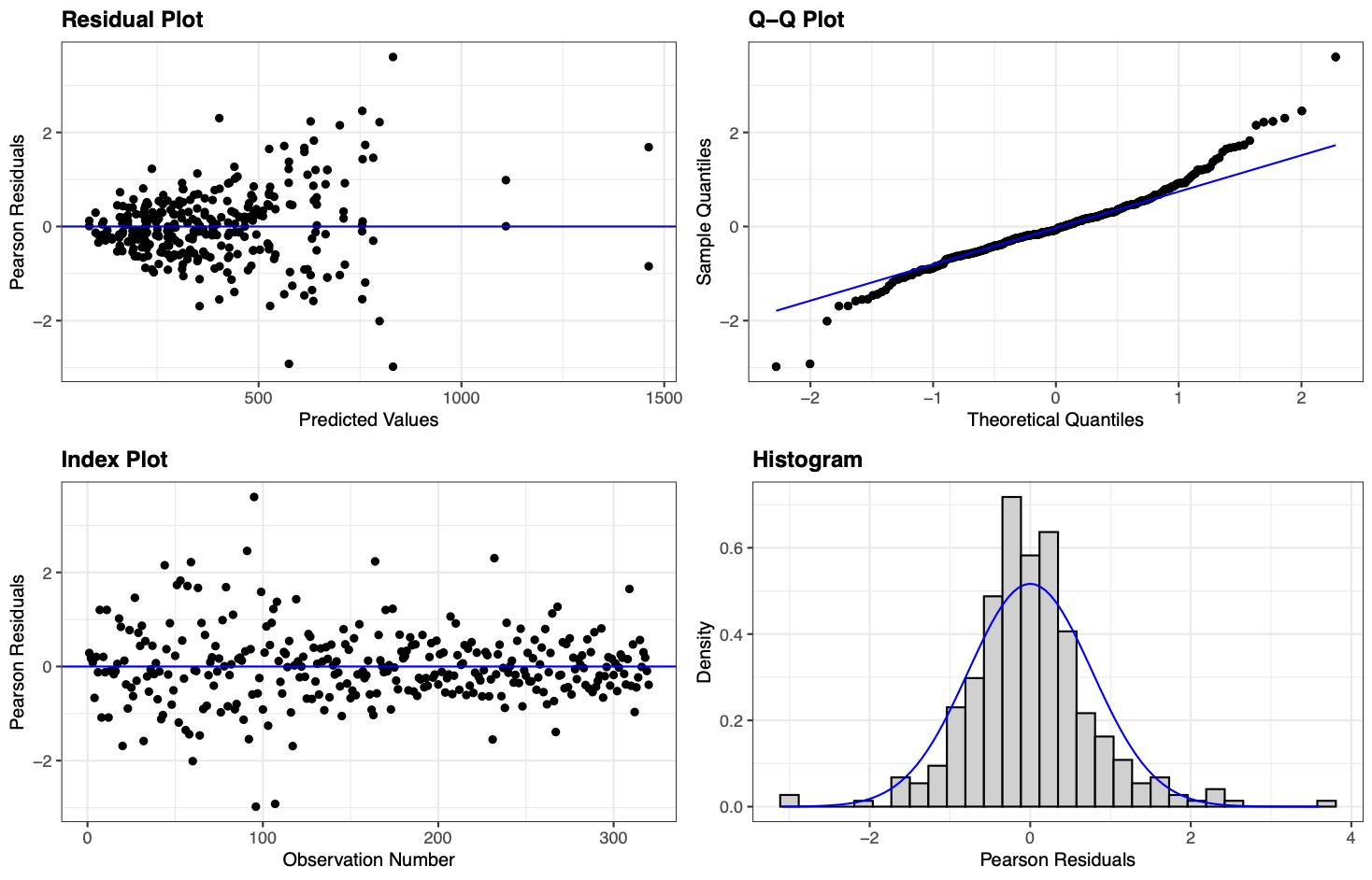

eTable 1. Conditioned pain modulation: model selection

| Model specification | Model name | Fixed Effects | Random Effects | Error/ Warning | Model fit | | |
| --- | --- | --- | --- | --- | --- | --- | --- |
|  |  |  |  |  | AIC/BIC | LL | df.resid |
| Linear model: pre-registered model | model1 | Condition*Group + Order | Condition + Order \| Subject | Model convergence problem; singular convergence | 3910.1/3955.3 | -1943.1 | 308 |
| Linear model: exclude order | model2 | Condition*Group | Condition \| Subject |  | 3916.6/3946.7 | -1950.3 | 312 |

eTable 2. Condition pain modulation: model estimates for the mixed effects model (model2)

| Fixed effects | | | | | |
| --- | --- | --- | --- | --- | --- |
|  | Est/Beta | SE | 95% CI | df, *t* | *P* |
| Intercept | 246.42 | 25.78 | 195.28 - 297.60 | 80, 9.56 | <0.001 |
| Condition_CS_ | 101.78 | 17.58 | 66.90 - 136.67 | 80, 5.79 | <0.001 |
| Group_NSSI_ | 124.21 | 36.01 | 55.77 - 195.65 | 80, 3.45 | <0.001 |
| Condition_CS_ × Group_NSSI_ | -94.32 | 24.56 | 45.59 - 143.04 | 80, 3.84 | <0.001 |
| Random effects | | | | | |
|  | Variance | SD | Correlation | | |
| Subject (intercept) | 24050 | 155.08 |  | | |
| Subject × Condition_CS_ (Slope) | 8309 | 91.15 | 0.10 | | |
| Residual | 3751 | 61.24 |  | | |
| *Note*. P-values for fixed effects were calculated using Satterthwaites approximations. Model formula: kPa ~ Condition * Group + (Condition \| Participant). N = 80, total observations = 320 | | | | | |

**Temporal summation**

The code that accompanies the results presented in eTable 3 can be found in ‘tempsum_results.R’ in the R-script folder on https://osf.io/gujwt/.

eTable 3. Temporal summation: model estimates for the mixed effects model

| Fixed effects | | | | | |
| --- | --- | --- | --- | --- | --- |
|  | Est/Beta | SE | 95% CI | df, *t* | *P* |
| Intercept | 0.87 | 0.09 | 0.69 - 1.05 | 76.61, 9.51 | <0.001 |
| Stimulus_max_ | 1.77 | 0.19 | 1.40 - 2.15 | 76.75, 9.34 | <0.001 |
| Group_NSSI_ | -0.05 | 0.13 | -0.31 - 0.22 | 76.19, -0.34 | 0.734 |
| Stimulus_max_ × Group_NSSI_ | 0.10 | 0.28 | -0.45 - 0.65 | 76.51, 0.36 | 0.723 |
| Random effects | | | | | |
|  | Variance | SD | Correlation | | |
| Subject (intercept) | 0.25 | 0.50 |  | | |
| Subject × Stimulus_max_(slope) | 1.28 | 1.13 | 0.38 | | |
| Residual | 0.30 | 0.55 |  | | |
| *Note*. P-values for fixed effects were calculated using Satterthwaites approximations.  Model formula: Rating ~ Stimulus * Group + (Stimulus \| Participant). | | | | | |

**fMRI**

When restricting the analysis to a priori defined pain regions, using the Neurologic Pain Signature (NPS) template, NSSI had higher activations in three clusters compared to controls: mid cingulum (x = 2, y = 6, z = 44, t = 4.02, *P* = .014), anterior cingulum (x=-5, y=21, z=30, t=3.58, p=.037), and the anterior insula (x = 40, y = 11, z = 2, t = 3.55, *P* =.037). There were no regions where HC had greater neural activations within the NPS than NSSI.

**Questionnaires**

Internal consistency for the questionnaires assessed by NSSI participants are presented in eTable 4.

eTable 4. Internal consistency for the questionnaires used in the NSSI group (n=41)

|  | Number of items | Cronbach’s α |
| --- | --- | --- |
| Borderline Symptom List | 23 | .95 |
| Self-Rating Scale | 8 | .82 |
| Difficulties with Emotion Regulation Scale | 16 | .93 |

**References**

1. Meteyard L, Davies RA. Best practice guidance for linear mixed-effects models in psychological science. *J Mem Lang*. 2020;112:104092. doi:https://doi.org/10.1016/j.jml.2020.104092.
